# COVID-19 apparent reproductive number dropped during Spain’s nationwide dropdown, then spiked at lower-incidence regions

**DOI:** 10.1101/2020.05.30.20117770

**Authors:** L. Santamaría, J. Hortal

## Abstract

COVID-19 pandemic has rapidly spread worldwide. Spain has suffered one of the largest nationwide bursts, particularly in the highly populated areas of Madrid and Barcelona (two of the five largest conurbations in Europe). We used segmented regression analyses to identify shifts in the evolution of the apparent reproductive number (Rt) reported for 16 Spanish administrative regions. We associate these breaking points with a timeline of key containment measures taken by national and regional governments, applying time lags for the time from contagion to case detection, with their associated errors. Results show an early decrease of Rt that preceded the nationwide lockdown; a generalized, sharp decrease in Rt associated with such lockdown; a low impact of the strengthened lockdown, with a flattening of Rt evolution in high-incidence regions regions – but increases in Rt at low-incidence regions; and an increase in Rt, associated to the relaxation of the lockdown measures, in ten regions. These results evidence the importance of generalized lockdown measures to contain COVID-19 spread; and the limited effect of the subsequent application of a stricter lockdown (restrictions to all non-essential economic activities). Most importantly, they highlight the importance of maintaining strong social distancing measures and strengthening public health control during lockdown de-escalation.

## Introduction

The rapid spread of COVID-19 pandemic has led to over 5.8 million confirmed cases from 31 December 2019 to 29 May 2020 (John Hopkins University Coronavirus Resource Center, see Dong et al. 2020). Such growth has been spatially heterogeneous, with large differences between countries, and between regions within countries. These differences were partly due to their different responses to the WHO alert, based on the resources and structure of their public health and R&D systems and their ability to implement social distance measures (see, e.g., Fernández-Recio 2020). But also to their international connectivity (Coelho et al. 2020), the specific strains reaching each territory (see updated data at nextstrain, Hadfield et al. 2018) and the age and spatial structure of their human populations (see Gatto et al. 2020). Response within countries has also been heterogeneous, particularly for those with decentralized governance structures – such as the federal states of USA, Germany and Brazil, or the administrative regions of Italy or Spain (see, e.g., Gatto et al. 2020).

COVID-19 arrived to mainland Spain at least in early February (first recorded hospitalization dates back to 15 February; Table S1), if not before (see Deslandes et al. 2020). During February, COVID-19 infection reached Spain repeatedly, mainly via UK, Italy and China – as evidenced by the presence of fourteen different genetic clusters identified by nextstrain (Hadfield et al., 2018; data from 27 April 2020). The combination of these repeated introductions with early, unnoticed community transmission resulted in consecutive outbreaks in distant, highly populated areas of the Basque Country and Navarra (North), Madrid (Center), Catalonia (North East), Andalusia (South) and Valencia (East). It is reasonable to assume that, by early March COVID-19, infections were widely distributed throughout the whole country. The pandemic peaked, however, most strongly in Madrid (around 6.4M people; second most populated metropolitan area of the EU, after Paris) – to the point that 65–67% of local incidence and mortality at the 52 Spanish provinces is explained by their early-stage mobility from and to Madrid (Mazzoli et al. 2020).

The adoption of containment measures by the national and regional governments followed a sustained increment through time, from (1) the recommendation of preventive measures in late February and early March, to (2) increasingly stricter social-distancing measures on 9–10 March, to (3) a nationwide lockdown announced on 13 March and enforced on 15 March, to (4) a strengthened lockdown, with the closure of all non-essential economic activities on 31 March, to (5) a cessation of the strengthened lockdown on 13 April, to (6) a relaxation of the lockdown measures, allowing daily outings by children (26 March), the elderly, and adults on sport and leisure activities (4 May), to (7) a de-escalation calendar for the progressive cessation of the lockdown, which started on 11 May (although its evolution is adjusted at province level based on incidence and medical criteria: some Balearic and Canarian Islands entered on 4 May, and Madrid, Catalonia, Castilla-León, Valencian Community and most of Castilla-La Mancha did not qualify yet to enter on 11 May).

In an earlier analysis of the cumulative numbers of cases and deaths – restricted to a period ending in mid-April 2020 due to the inconsistencies in official data after such date, we characterized the impact of governmental restrictions on mobility and personal contact at three spatial levels: the whole of Spain, and Madrid and Catalonia Autonomous Regions (Santamaría & Hortal 2020)). Our results showed an early deceleration in the spread of COVID-19, coinciding with personal hygiene and social distancing recommendations, and the effectiveness of enforcing harder isolation measures to contain the disease outbreak by early April. In this paper, we use official daily estimates of the apparent reproductive number (Rt) at regional level to evaluate the responses of the pandemic to early containment measures (social distancing and hygiene, nationwide lockdown, and strengthened lockdown) and their subsequent relaxation.

## Data and methods

### Infection and fatality data

Official data were obtained from the centralized panel on the “Situation and evolution on the COVID-19 pandemic in Spain” run by the National Epidemology Center (Instituto de Salud Carlos III – CNE-ISCIII) and the Spanish Ministry of Health. Daily estimates of the apparent reproductive number (Rt) aggregated at regional level (autonomous regions) were extracted from the site (Evolution of the pandemic > Aggregated declaration) for all Spanish Autonomous Communities (regions hereafter) except the Canary Islands, Ceuta and Melilla (which were excluded because of their small size and/or isolated geographic location). Estimates are based on cumulative data (PCR-confirmed cases until the day of reporting) provided by the regional governments to the Ministry of Health; and calculated with R EpiEstim statistical package, using a Poisson model and a Bayesian framework to construct confidence intervals, and a seven-day aggregation window (as reported in the site’s ‘Documentation and data’ page). The data series has been discontinued on 20 May 2020, owing to changes in the reporting procedures associated to the new Vigilance and Control Strategy – which require a re-consolidation of existing data before the addition of new ones.

The time series used for the analysis ranges from 14 March 2020 –the first date for which data were available for all regions, and 20 May 2020 –when data updating was temporarily halted (see above). Hence, it starts the day in which the nationwide lockdown was declared, and finishes nine days after the onset of the de-escalation. We used the official estimates of Rt, instead of our own calculations based on reported numbers of cumulative cases, because (i) continued update of the latter (to correct for disparities in the reporting times and procedures of the different regional authorities) by the central authority, without reporting clearly on the procedures or changes made, makes the former more reliable; and (ii) because these are the data (putatively) used for decision making during the preceding and forthcoming periods.

### Lag time estimates

We followed Santamaría & Hortal (2020) to estimate the infection date of reported cases. Briefly, we calculated the infection-to-testing time by combining the reported values of incubation time (mean = 5.0 days from Lauer et al., 2020; median = 5.1 days from Linton et al. 2020; mean = 6.4 days from Lai et al. 2020) with time from illness onset to hospital admission for treatment and/or isolation (median = 3.3 days among living cases and 6.5 days among deceased; Linton et al. 2020). Hence, we used an infection-to-testing time of 9 days for living cases and 12 days for dead cases. Based on the proportion of 36% deaths to 64% recoveries reported from 3 March 2020 to 6 April 2020 (for a total of 57,006 closed cases) in Spain, we estimated an average infection-to-testing time of 10.1 days – which, for simplicity, was rounded to 10 days.

### Analyses

We used segmented regressions (i.e. broken-line) to identify breakpoints in the temporal behavior of apparent transmission rate values in the 16 Spanish regions analyzed. Changes in the slope of daily Rt values are taken to represent shifts in COVID-19 transmission trends. We fitted a family of segmented regressions ranging from no to eight breaking points, with a minimum of five data points per segment, and compared them using their respective BICs, using the strucchange (breakpoints function; Zeileis et al. 2001) and lme4 (for the calculation of the best-model’s intercepts and slopes) packages of R 3.6.3 (R Core Team 2020). The algorithm of strucchange’s breakpoints use separate intercepts at each different segment, thus allowing for discontinuities in the fitted functions – and thus for the separate identification of progressive increases or decreases in Rt, and sudden “jumps” or “dives” in daily values of this parameter.

## Results

The segmented models fitted successfully all regional data series (Figure 1). The number of breakpoints ranged from only two (in Castilla-León and the Basque Country) to a maximum of eight (in Asturias). Note that, hereafter, references to the association between breakpoints and management measures will already apply the expected 10-day delay between infection and detection (as indicated in Fig. 1’s upper X-axis arrows).

In most regions (14 out of 16), decreases in Rt preceded the onset of the nationwide lockdown (albeit with a considerable delay for Balearic Islands). Indeed, in Madrid, Catalunya and Castilla-León, the drop in Rt that started before the national lockdown simply continued at the same rate after it. In contrast, in the Basque Country and La Rioja Rt kept increasing until 26–27 March and showed a turning point coinciding with the enforcement of the nationwide lockdown. Moreover, in nine more regions (most notably Aragón, but also Castilla-La Mancha, Andalusia, Valencia, Galicia, Navarra, Extremadura, Cantabria and Murcia), the decrease in Rt previous to the nationwide lockdown was interrupted by increases or upward jumps in Rt values (see Figure 1).

**Figure 1:**
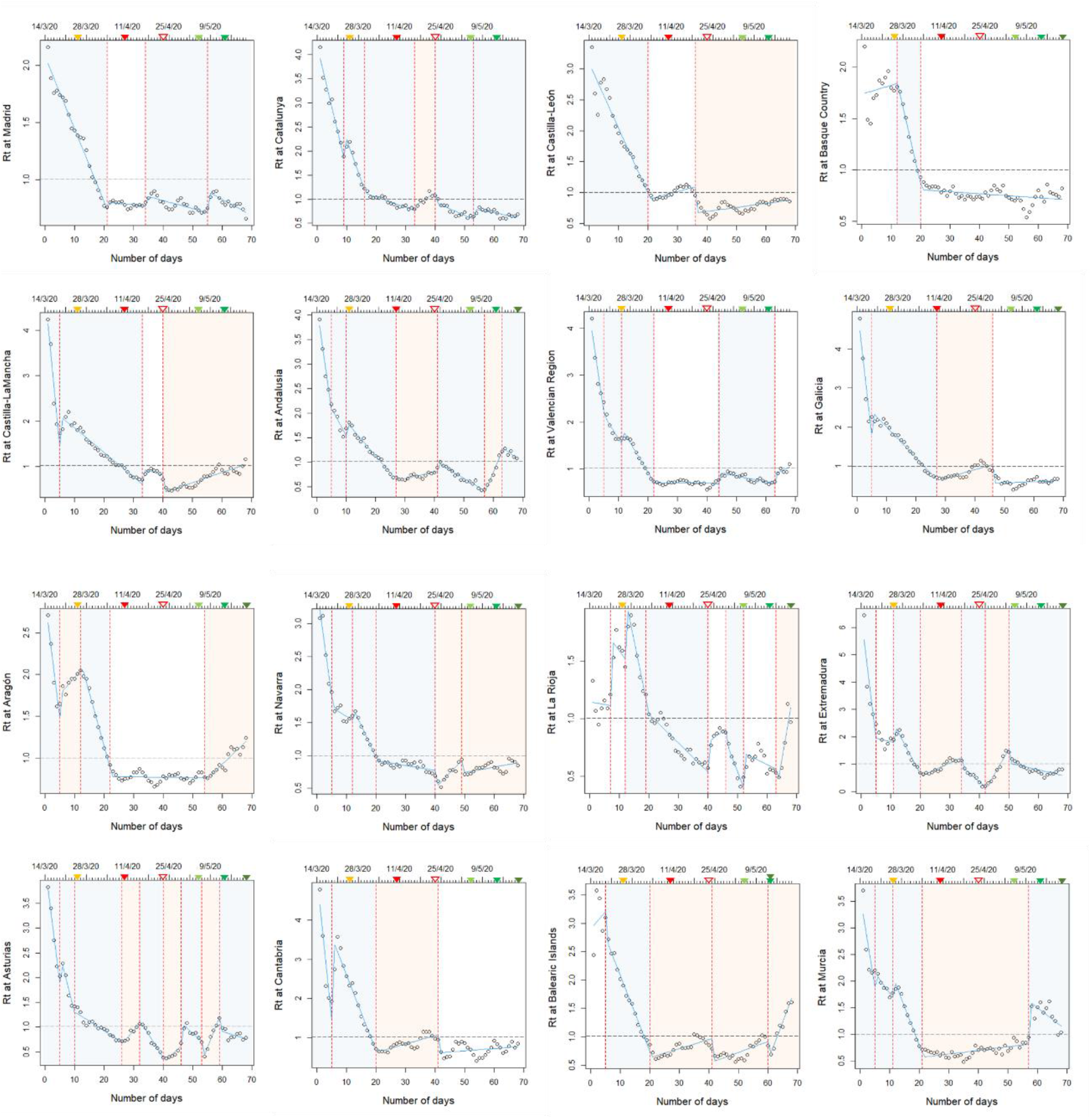
Results of segmented regressions on the apparent reproductive number (Rt) of the COVID19 pandemic in the different autonomous regions of Spain. In the upper X axes (dates), colored triangles indicate the estimated detection times of infections taking place at the onset of key policy measures: orange = nationwide lockdown (13 March); filled red = strengthened lockdown (31 March); empty red = end of the strengthened lockdown (13 April); light and medium green = relaxation of lockdown for children (26 March) and elderly/adults (4 May), respectively; dark green = phase 1 of de-escalation program (variable among regions). Red broken lines indicate breakpoints. Blue and red areas respectively indicate segments with significant increases and decreases in Rt.

The enforcement of the nationwide lockdown marks the strongest and most widespread turning point in the whole dataset. In all regions, the lockdown resulted in a swift decrease in Rt until reaching values < 1 (indicating the switch from the spread to the control phases of infection dynamics) and stabilizing there. In contrast, the subsequent enforcement of the strengthened lockdown (cessation of all non-essential economic activity) had limited to no impact on the containment of the epidemic: it resulted in no additional decreases in Rt, and was associated instead to switches to positive slopes (Rt increases) in at least five regions (Andalusia, Galicia, Cantabria, Balearic Islands and Murcia). In general, the period between the onset and the end of the strengthened lockdown was characterized by low Rt values (below or slightly above 1) and either flat or fluctuating trends.

The end of the strengthened lockdown was closely associated with major changes in the infection trends, with long term increases in Rt in at least four regions (Castilla-León, Castilla-La Mancha, Navarra and Balearic Islands), shorter-term increases in two more (Extremadura and Asturias) and a large ‘jump’ in values in yet another one (La Rioja).

Finally, the de-escalation coincided with sudden and sustained increases in Rt in seven regions (Andalusia, Valencia, Aragón, La Rioja, Asturias, Balearic Islands and Murcia), and prolonged the increasing trends initiated after the relaxation of the strengthened lockdown in three more (Castilla-León, Castilla-La Mancha and Navarra).

Strikingly, the likelihood of switching to an increase in Rt values was higher in regions that were hit less hard by the pandemic. There was a significant, negative relationship between the incidence of the pandemic (rank based on the number of cases per region) and the occurrence of Rt increases during the strengthened confinement (logistic regression: χ^2^ = 4.73, df = 1, P< 0.030, logistic regression with probit link; Figure 2), but not during the relaxation (end of strengthened lockdown) and de-escalation periods (χ^2^ = 0.58, df = 1, P>0.44 and χ^2^ = 1.36, df = 1, P>0.24, respectively).

**Figure 2:**
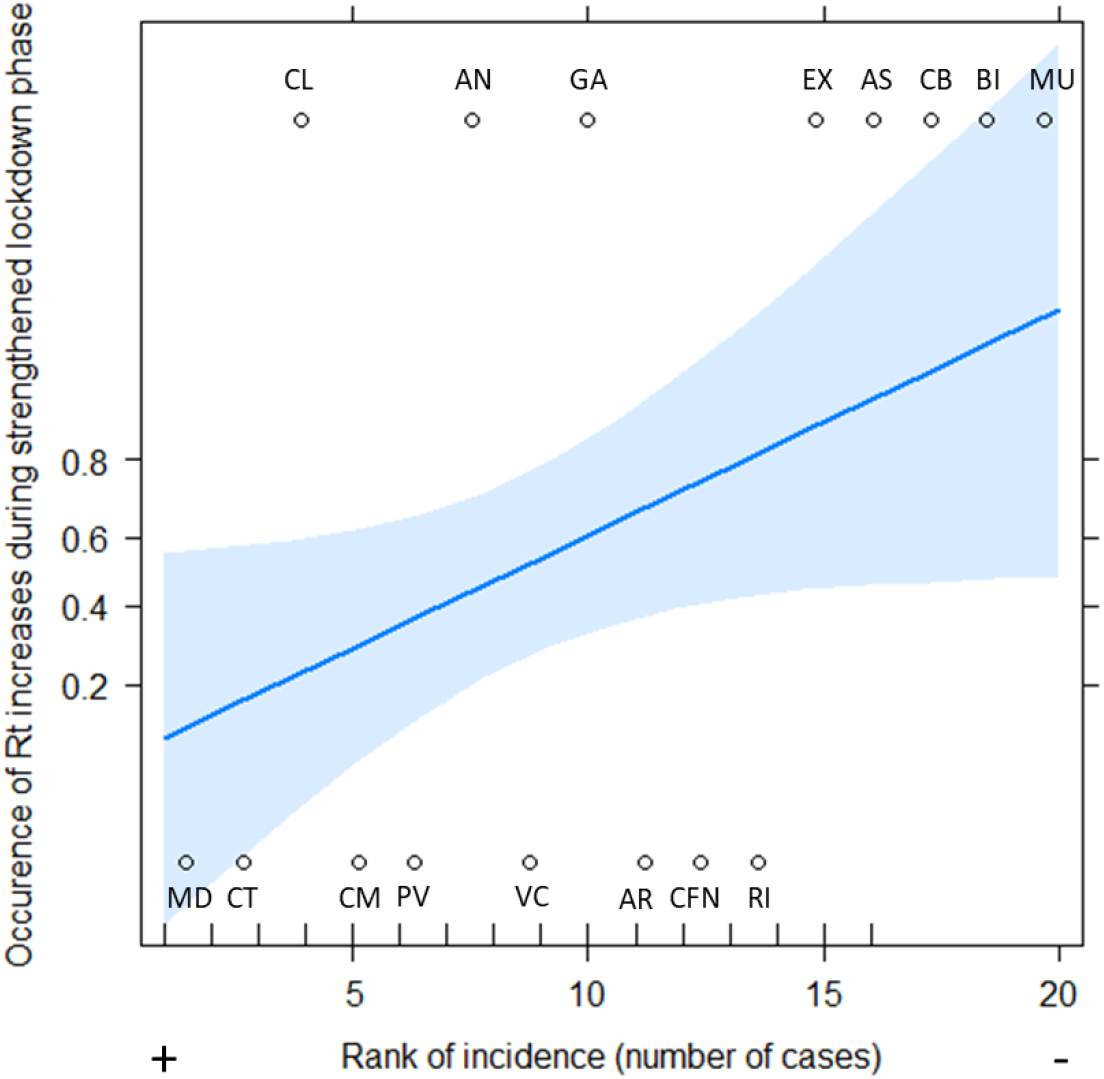
Relationship between the occurrence of increases in apparent reproductive number (Rt) and the rank of incidence (from higher to lower number of cases) in each Spanish region.

## Discussion

Our results show an early deceleration in the spread of COVID-19, probably resulting from personal hygiene and social distancing recommendations that preceded the enforcement of the nationwide lockdown in most regions (see also Santamaría & Hortal 2020). The nationwide lockdown marks a clear turning point in the apparent reproductive number: the strongest decrease in Rt detected in all regions follows such enforcement (see also Tobías 2020). However, in nine regions the decrease in Rt that preceded the nationwide lockdown was interrupted by increases in Rt values (most notably Aragón, Castilla-La Mancha, Navarra and Extremadura). It is likely that these increments are related to the increases in inter-regional mobility from the most affected regions (Madrid, Catalonia, Basque Country and La Rioja; see Maxxoli et al. 2020), triggered by local lockdown measures (such as the closure of schools and universities in Madrid and parts of Basque Country, in March 11) that unwittingly incentivized the migration of students and wealthy citizens to family homes and second residences (a response observed also in other countries, such as France and the US; Onishi and Méheut, 2020; Hoffower 2020).

In contrast with this strong effect, the strengthened lockdown did not result in further Rt decreases in any region. Instead, it resulted in counter-intuitive increases of transmission rate in several regions – with the four clearest examples involving coastal regions: Cantabria, Andalusia, Galicia and Balearic Islands. These responses may be the result of the relaxation and tiredness following several weeks of demanding lockdown – since they took place predominantly in communities with a lower perception of risk, resulting from a lower incidence of the pandemic (Figure 2; see also CISNS & ISCIII 2020); perhaps combined with an increased arrival of tourists during the Easter holidays – since they took place primarily in regions that represent major tourist destinations. This is best exemplified by two regions, Balearic Islands and Murcia, which had the lowest number of cases within the dataset – and showed a concatenation of increases in Rt from the beginning of the strengthened lockdown and through the relaxation and de-escalation periods, until the end of the time series (Figures 1 and 2).

The strengthened lockdown may have been instrumental, however, in maintaining the preventive tension and halting the recurrent spikes in Rt values – since its cessation was closely associated to major changes of trend, with long term increases in Rt in at least four regions (Castilla-León, Castilla-La Mancha, Navarra and Balearic Islands), shorter-term increases in two more (Extremadura and Asturias) and a large ‘jump’ in values in yet another one (La Rioja). As before, these changes may be related to increased inter-regional mobility – which, until then, was only authorized for transportation of essential goods and specialized personnel.

Importantly, the onset of the two de-escalation steps included in the time series seemingly caused sudden and sustained Rt increases in in seven regions, and prolonged the increasing trends initiated after the relaxation of the strengthened lockdown in three other ones. These responses are not significantly related to the incidence of the pandemic until that date, and seem instead to reflect a variability of causes across the different regions. For example, the increase during strengthened lockdown followed by a decrease during its relaxation in two regions (Andalusia and Galicia) is suggestive of a displacement of workers returned from and reincorporated to industries placed at larger cities, such as Madrid (see Mazzoli et al. 2020). Further research on mobility trends will be needed to elucidate the causes of the contrasting responses of the different Spanish regions.

Given the high basal level of community transmission in most of Spain and its surrounding countries, these increases can be easily triggered by single contagion events at social gatherings. For example, in the last week of May, four holiday parties of around 20, 80, 15 and 30 attendants resulted in outbreaks at Lleida (Catalonia), the autonomous city of Ceuta, Badajoz (Extremadura) and Córdoba (Andalusia). (The last example involved an infected Belgian resident who travel internationally to attend the party, despite all the restrictions in place.)

In summary, the swift decrease in Rt triggered by the national lockdown in all regions, was followed by: (i) moderate increases in Rt during the strengthened lockdown in c. half of the regions; and (ii) stronger upward spikes during the early relaxation and de-confinement phases. These increases brought Rt back from values well below zero to values above such threshold, and thus represent a tangible risk of entering a second infection wave. These circumstances seem most inadequate to introduce changes in the monitoring and reporting procedures, which have caused a halt in official data availability. A positive lesson may however be drawn from the early (pre-lockdown) decrease of Rt, caused primarily by voluntary hygiene and social distancing measures (Santamaría & Hortal 2020). While economic pressures may have pressured the authorities into committing to an early de-escalation, responsible citizen behavior may still contribute significantly to contain the risk of a second wave.

## Data Availability

All data were obtained from and are thus available at data repositories referenced in the Methods section

